# Knowledge, attitudes and practices toward female genital schistosomiasis among community women and healthcare professionals in Kimpese region, Democratic Republic of Congo

**DOI:** 10.1101/2023.07.17.23292750

**Authors:** Cecilia Wangari Wambui, Joule Madinga, Mercy Gloria Ashepet, Maxson Kenneth Anyolitho, Patrick Mitashi, Tine Huyse

## Abstract

**Background:** Chronic infection with *Schistosoma haematobium* causes female genital schistosomiasis (FGS), which leads to diverse lesions in the female genital tract and several complications, including women infertility and a higher risk for HIV transmission. This study, therefore, aims to understand the knowledge, attitudes, and practices (KAP) toward FGS and associated factors among women and health professionals in the schistosomiasis endemic focus of Kimpese, western Democratic Republic of Congo (DRC).

**Methods:** In January 2022, a semi-quantitative questionnaire was administered to randomly selected community women in Kifua II village and health professionals (nurses and doctors) from Kimpese Health Zone. KAP statements were coded and summarized as frequencies and percentages. Association between the socio-demographic characteristics of respondents and KAP was assessed using Pearson chi-square (χ2) test, Cramer’s V (φ) and gamma (γ) coefficients.

**Results:** A total of 262 participants were included (201 community women, 20 nurses and 41 doctors). Overall, respondents had high knowledge of schistosomiasis in general but low FGS- specific knowledge (91% versus 45%). Some misconceptions regarding FGS transmission were even higher among healthcare professionals compared to the community women. Almost a third (30%) of the community women and 20% of the nurses believe that FGS is transmitted by drinking untreated water, while 27% of the doctors believe that sexual contact is a mode of FGS transmission. Additionally, 30% of the doctors do not link FGS with urinating in the water. Furthermore, many community women (60%) practice open defecation or urination and do not consider avoiding contact with contaminated water sources important (72%), especially the younger ones. Finally, diagnostic technologies for FGS are lacking, with only 57% of healthcare workers having a microscope in their facilities.

**Conclusion:** This study reveals insufficient knowledge about FGS and existing negative attitudes toward FGS among community women associated with socio-demographic factors. Additionally, health professionals lack the means (equipment) and specialised knowledge to diagnose FGS correctly, which probably leads to underreporting as this region is endemic for urinary schistosomiasis.

**AUTHOR’S SUMMARY:** Schistosomiasis is a disease caused by parasitic worms and it is contracted through contact with contaminated water. Female genital schistosomiasis (FGS) is a form of the disease that affects the female reproductive organs and can lead to infertility, but also stigma and discrimination. This study used a quantitative research approach to explore community women and healthcare workers’ knowledge, attitudes and practices on and health-seeking behaviour of the women regarding FGS in Kongo Central, DRC. While both groups had high knowledge of schistosomiasis, they had a limited understanding of FGS specifically. Misconceptions about its cause and prevention were common, particularly among medical doctors. Many community members engaged in risky water contact and hygiene practices. Healthcare workers reported limited diagnostic tools and expressed interest in specialised training on FGS diagnosis, treatment, and prevention. Consequent to limited knowledge and ill-equipped laboratories, health workers are most likely underreporting the disease in the region. The study highlights the need to integrate FGS into public health education programs for both community members and healthcare workers in Kongo Central and underscores the importance of addressing misconceptions about the disease and the prevention measures.

## 1. INTRODUCTION

Human schistosomiasis (generally known as bilharzia or snail fever) is a neglected tropical parasitic infection [1], endemic to communities living in areas that have no access to adequate safe water and sanitation amenities [2]. There are over 240 million schistosomiasis infections globally, with 95% of these prevailing in Africa [2–5]. Schistosomiasis ranks second to malaria in terms of prevalence, constantly raising public health and socio-economic apprehension in Africa [6]. Three main species affect humans on the African continent: *Schistosoma haematobium* causes urogenital schistosomiasis, while *Schistosoma mansoni* and *Schistosoma intercalatum* cause intestinal schistosomiasis. The respective parasite species are transmitted through freshwater snails that belong to different genera, namely *Biomphalaria* (*S. mansoni*) and *Bulinus* (*S. intercalatum* and *S. haematobium*). When a person gets into contact with infested water, the parasite larvae (cercariae) penetrate the skin [7,8]. Both forms of schistosomiasis (intestinal and urogenital) cause anaemia, stunted growth, and reduced learning ability in children. The eggs of *S. mansoni* and *S. intercalatum* are lodged in the mesenteric veins of the intestines, while those of *S. haematobium* are lodged in the veins surrounding the urogenital system [9–11]. The parasite eggs trigger an inflammatory immune response that causes acute or chronic illness. The disease is often not fatal, but in its chronic form, *S. mansoni* and *S. intercalatum* mainly lead to abdominal pain, bloody diarrhoea, severe fever, hepatomegaly and hepatic fibrosis, while chronic *S. haematobium* infection can lead to haematuria, irritation in the bladder, bladder cancer, painful and frequent urination [1].

Chronic infection with *S. haematobium* also causes female genital schistosomiasis (FGS), characterised by schistosome eggs and /or a distinctive pathology in the female reproductive system [12–14]. Eggs that are trapped in the urogenital tissues will provoke an inflammatory reaction with diverse lesions in the genital tract of women and girls [12–14]. Other symptoms include postcoital haemorrhage, itchiness, unusual discharge, dyspareunia and incontinence [9,15]. Additionally, non-specific symptoms, for example, vaginal discharge and itchiness, are often mistaken for sexually transmitted illnesses. This leaves many girls and women misdiagnosed, leading to the affected females, especially young girls, to be accused of sexual promiscuity, which in turn causes stigma and discrimination [16]. Other ripple effects associated with FGS include marital conflicts and emotional distress [16]. Besides these adverse reproductive, emotional and social consequences, FGS is also known to be a co- factor in acquiring human immunodeficiency virus type 1 (HIV-1), human papillomavirus (HPV) and cervical cancer [14,17–19].

Schistosomiasis genital tract manifestation has been neglected by academia compared to urinary and intestinal manifestation in terms of the number of publications [19]. In addition, the risk to sexual reproductive health is neither sufficiently addressed in public health interventions, nor are the prevention strategies being pursued [9,20]. FGS is often linked to problematic detection, as diagnosis requires specialised equipment (colposcope) and training, which leads to underreporting in most endemic areas, hence, a lack of reliable data on prevalence. However, based on the prevalence of *S. haematobium*, an estimated 56 million women and girls in most communities of Africa are at risk of FGS infection [16]. Moreover, apart from the communities lacking knowledge of FGS, many healthcare professionals are unaware of the disease, diagnosis, risks, and treatment [16]. Therefore, increasing awareness among the communities and healthcare professionals is important to enable early detection of FGS and focus on preventive measures. Studies show that community mobilization and participation (bottom-up approach) are key to sustainable and compelling communication to inform the affected communities on symptoms, prevention and control of FGS [21,22]. However, for bottom-up approaches to be successful, the existing knowledge, attitudes and practices (KAP) toward the disease need to be assessed [23]. The KAP information gathered would inform the development of effective control strategies. This study, therefore, sought to fill this gap by assessing the KAP toward FGS and sociodemographic characteristics associated with the KAP among community women and health professionals in a highly endemic region in Kongo Central province, DRC.

## 2. METHODOLOGY

### 2.1 Ethical considerations

The study was approved by the Ethical Committee of the University of Kinshasa, DRC (n°191/CNES/BN/PMMF/2020) (SI1). Before the start of the interviews, the community members were invited on the 7^th^ of January 2022 for a sensitization meeting at Kifua II local health centre, where the village chief and the nurses of Kifua II local health centre were present. Permission to interview the medical doctors and the nurses was sought from the Chief Medical Officer at Kimpese Health Zone. The community surveys were conducted at the women’s households in a comfortable private setting of their choice. The doctors were visited at their respective working facilities in Kimpese city, while nurses were invited at the Kimpese Health Zone office. Facemasks were provided before the start of the interviews, given the COVID-19 pandemic context. The interviewer explained the aims of the study and the guaranteed confidentiality of the data collected to the participants. In addition, they were informed about the right to stop their participation at any time without any consequence and were encouraged to ask questions about the research. Also, verbal informed consent was obtained from each respondent before participating in the study and parents consented on behalf of the girls below 18 years.

### 2.2 Study area and setting

This study was conducted in January 2022 in two areas of Kongo Central province of DRC: **(i)** community women were interviewed in Kifua II village located in the Health Zone (HZ) of Kwilu Ngongo and **(ii)** the medical doctors and nurses in the city of Kimpese HZ (Fig 1). Kongo Central province is known to be endemic for urogenital schistosomiasis [24,25] and Kifua II was selected based on its high prevalence (68%) of schistosomiasis infections and the recent studies on reinfection morbidity and co-infection of schistosomiasis with salmonella [25], Madinga, unpublished]. Kifua II lies at 20 Km from Kimpese on the national road n°1 and borders the Ngongo River, which supplies water to many households in the community. The city of Kimpese includes the central office of the rural HZ of Kimpese and the general referral hospital of this zone. Kimpese HZ is characterised by poor hygiene, with previous studies reporting 41% not having a toilet in the household, while 48% opting for open defecation and urination [26]. Furthermore, there is no prior record of mass drug administration (MDA) for schistosomiasis control in Kifua II village [27] [24]. However, treatment was provided to the infected people after the schistosomiasis prevalence study in 2015 [Madinga, unpublished].

**Fig 1.**
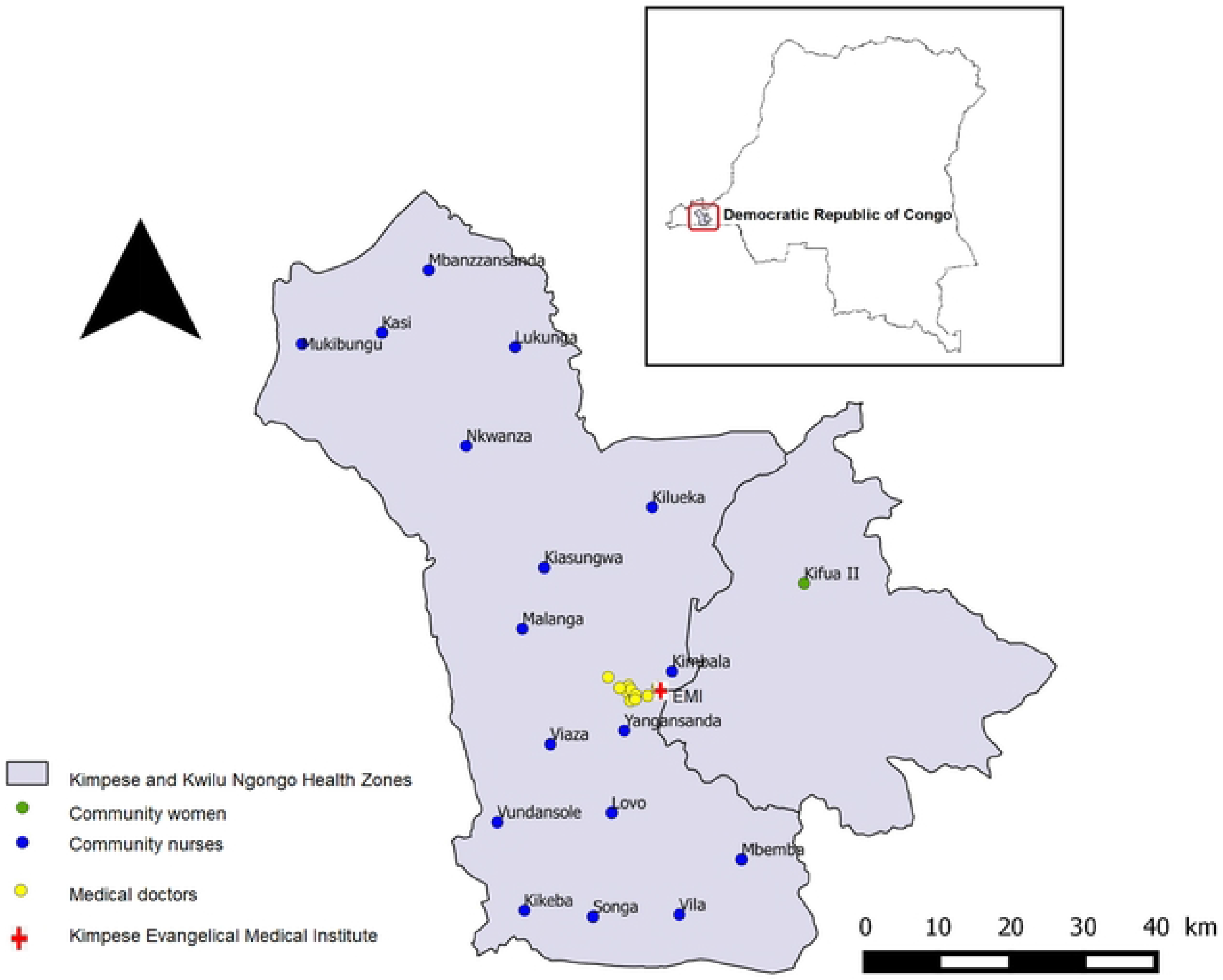
Study area showing locations of the study groups in Kongo Central, DRC.

### 2.3 Study design

A cross-sectional study design was employed with two semi-structured questionnaires for three weeks in January 2022: 1) for the community women 2) another for the healthcare professionals; with both open-ended and closed-ended questions adapted from Anyolitho et al. and Masong et al. [28,29]. The questionnaires were translated from English to French before feeding them into the open-source data collection application “KoBoToolbox”. The application enables researchers to collect data offline in remote settings and later upload the gathered information to a server once connected online [30]. The translated versions were administered by 8 trained field researchers that were competent in both French and the local language (Kikongo); six of them were female (two from Kifua II and four from Kimpese) and two were male (one of them was a medical doctor from Kimpese). The two target study groups comprised of the community women in Kifua II (only interviewed by the female researchers) and healthcare professionals (nurses and doctors) in Kimpese (all the doctors were interviewed by the medical doctor interviewer).

The questionnaire (Supplementary Information – SI2-3) was subdivided into four sections: **(i)** demographic information; **(ii)** knowledge indicators or familiarity with the signs and symptoms, modes of transmission, control methods and risk factors; **(iii)** attitudes toward the disease and perceptions toward the infected individuals (state of stigmatization) (statements for the community women were different from those of the healthcare professionals); **(iv)** socio- cultural and management practices related to the spread of FGS and the health-seeking behaviour of the community women. Information guides were provided to the community women and the healthcare professionals after the interviews, with relevant information about FGS (SI4a-4b). The information guides were made based on available FGS literature and educational resources, such as from World Health Organisation (WHO), and then translated into French and Kikongo.

### 2.4 Selection and sample size determination

Sample size estimation for the women was based on similar studies conducted in Cameroon and Zimbabwe [27, 28]. The population size (N=431) of women in the target reproductive age was derived based on <u>United Nations World Population Prospects Report guidelines</u>, using the total population of Kifua II village, estimated to be 1,400 [24]. The report suggests that 49.6% of the world’s population is female and 62% of them are between the ages of 15 - 59 years. Hence, N = 1400 × 0.496 × 0.62. The sample size (n=203) was then estimated following Cochran’s formula n = [(z^2^pq)/e^2^]/[1+(z^2^pq/e^2^N)]; where, ‘n’ is the sample size, ‘Z’ is the critical value at 95% confidence level (1.96), ‘e’ is the margin of error (5%), ‘p’ is the sample proportion (50%), ‘q’ is 1-p and ‘N’ is the population size (431). The community women were selected through simple random sampling, while the selection of the healthcare professionals was purpose-driven. Head nurses from all the 20 local health posts in Kimpese Health Zone were chosen, while 41 of the available 44 medical doctors in Kimpese HZ participated in the survey.

### 2.6 Data processing and statistical analysis

The independent variables were the socio-demographic characteristics (age, level of education, monthly income, marital status and type of occupation). For the dependent variables, we used Likert scales for knowledge, attitude and practices. The knowledge score was obtained by combining all the correct answers (coded one) given by the respondents. Total scores (maximum of 25 correct answers) were a combination of the knowledge of symptoms, complications and modes of transmission, with low knowledge (coded zero) ranging from 0 to 8, moderate knowledge (coded one) ranging from 9 to 16 and high knowledge (coded two) ranging from 17 to 25. Statements to measure attitude were collected using a 5-point Likert scale ranging from 1 (strongly disagree) to 5 (strongly agree). Scores were computed by averaging 13 statements, with the high scores indicating positive attitudes towards the disease. Cronbach’s alpha (α) coefficient was used to assess the internal reliability and consistency of the Likert scales. The coefficient ranges from 0 to 1, with “_> .9 - Excellent, _> .8 - Good, _> .7 - Acceptable, _> .6 - Questionable, _> .5 - Poor, _< .5 - Unacceptable” [31]. For the practice section, the responses indicating good practice (not involved in activities that could lead to the spread or transmission of FGS; seeking medical attention when experiencing FGS-like symptoms) were coded one and zero for otherwise (responses that reflect bad practice). Data were analysed using IBM SPSS software version 19. Summarised descriptive statistics (frequency values and percentages) were obtained for the KAP variables and the respondents’ sociodemographic profile. Contingency tables were computed and Pearson chi-square (χ2) (for the nominal variables), Cramer’s V (φ) and gamma (γ) coefficients (for the ordinal variables) were recorded to ascertain the sociodemographic characteristics that are associated with the knowledge, attitudes, and practice variables among the respondents. For the interpretation of the results, the level of statistical significance was set at a *p*-value of 0.05, while the strength of the associations (using φ and γ coefficients) were as follows: >0.5 high association; 0.3 to 0.5 moderate; 0.1 to 0.3 low association; 0.0 to 0.1 little if any association [29].

## 3. RESULTS

### 3.1 Sociodemographic characteristics of the participants

A total of 262 participants were included (201 community women, 20 nurses and 41 doctors). The age of community women ranged between 15-59 years, with more than half (56.2%) aged 36 and below, while only 20.9% were older than 48 years. Most (65.6%) of them were married and over a third (39.8%) of the respondents never obtained primary education. The majority (59.7%) were farmers, while slightly over one-fifth (20.9%) were unemployed. Nearly two-thirds (60.2%) had monthly earning of less than 100,000 Congolese Dollar Franc (CDF) (equivalent to USD 50). Almost two-thirds (62.3%) of the healthcare professionals were between 29 and 45 years, with 85.2% of them being male. The majority (91.8%) have been practising for less than 20 years (SI5).

### 3.2 Knowledge of FGS

#### 3.2.1 Community women’s knowledge

Frequency of the level of knowledge by demographic characteristics of community women are given in the Table 1. Overall, more than half (55.7%) of women had a low level of knowledge regarding FGS. This trend was the same for all categories except among women with monthly income of 100,000 – 199,000CDF where 66.7% had a moderate level of knowledge.

**Table 1.** Knowledge, attitude and practices towards female genital schistosomiasis by socio-demographic characteristics of community women in Kifua II village.

| Socio-demographic characteristics | n | Knowledge (levels) |  |  | Attitudes (means) |  |  |  |  | Practices |  |
| --- | --- | --- | --- | --- | --- | --- | --- | --- | --- | --- | --- |
|  |  | Low | Moderate | High | 1 | 2 | 3 | 4 | 5 | Bad | Good |
| **All** | 201 | 112<br>(55.7%) | 80 (39.8%) | 9 (4.5%) | 70<br>(34.8%) | 11<br>(5.5%) | 3<br>(1.5%) | 98<br>(48.8%) | 19<br>(9.4%) | 5 (2.5%) | 196 (97.5%) |
| **Age (years)** |  |  |  |  |  |  |  |  |  |  |  |
| 15 - 25 | 61 | 38 (62.3%) | 22 (36.1%) | 1 (1.6%) | 22 (36%) | 7<br>(11.5%) | 2<br>(3.3%) | 27<br>(44.3%) | 3<br>(4.9%) | 1 (1.6%) | 60 (98.4%) |
| 26 - 36 | 52 | 29 (55.8%) | 21 (40.4%) | 2 (3.8%) | **27**<br>**(51.9%)** | 3<br>(5.8%) | 0 (0%) | 17<br>(32.7%) | 5<br>(9.6%) | 2 (3.8%) | 50 (96.2%) |
| 37 - 47 | 46 | 23 (50%) | 21<br>(45.65%) | 2<br>(4.35%) | 9<br>(19.6%) | 0 (0%) | 0 (0%) | 32<br>(69.6%) | 5<br>(10.9%) | 1 (2.2%) | 45 (97.8%) |
| 48 - 59 | 42 | 22 (52.4%) | 16 (38.1%) | 4 (9.5%) | 12<br>(28.6%) | 1<br>(2.4%) | 1<br>(2.4%) | 22<br>(52.4%) | 6<br>(14.3%) | 1 (2.4%) | 41 (97.6%) |
| **Education Level** |  |  |  |  |  |  |  |  |  |  |  |
| Without education | 80 | 43<br>(53.75%) | 33<br>(41.25%) | 4 (5%) | 24 (30%) | 4 (5%) | 0 (0%) | 45<br>(56.2%) | 7<br>(8.8%) | 2 (2.5%) | 78 (97.5%) |
| Primary Education | 108 | 61 (56.5%) | 43 (39.8%) | 4 (3.7%) | 39<br>(36.1%) | 7<br>(6.5%) | 3<br>(2.8%) | 49<br>(45.4%) | 10<br>(9.2%) | 3 (2.8%) | 105 (97.2%) |
| Secondary & high Education | 13 | 8 (61.5%) | 4 (30.8%) | 1(7.7%) | **5**<br>**(45.4%)** | 0 (0%) | 0 (0%) | 4<br>(36.4%) | 2<br>(18.2%) | 0 (0%) | 13 (100%) |
| **Marital Status** |  |  |  |  |  |  |  |  |  |  |  |
| Not married | 69 | 35 (50.7%) | 32 (46.4%) | 2 (2.9%) | 29 (42%) | 1<br>(1.4%) | 1<br>(1.4%) | 32<br>(46.4%) | 6<br>(8.7%) | 2 (2.9%) | 67 (97.1%) |
| Married | 132 | 77 (58.3%) | 48 (36.4%) | 7 (5.3%) | 41 (31%) | 10<br>(7.6%) | 2<br>(1.5%) | 66 (50%) | 13<br>(9.8%) | 3 (2.3%) | 129 (97.7%) |
| **Employment Status** |  |  |  |  |  |  |  |  |  |  |  |
| Unemployed | 100 | 58 (58%) | 40 (40%) | 2 (2%) | **43 (43%)** | 4 (4%) | 3 (3%) | 38 (38%) | 12<br>(12%) | 4 (4%) | 96 (96%) |
| Employed | 101 | 54 (53.5%) | 40 (39.6%) | 7 (6.9%) | 27<br>(26.7%) | 7<br>(6.9%) | 0 (0%) | 60<br>(59.4%) | 7<br>(6.9%) | 1 (1%) | 100 (99%) |
| **Monthly Income (CDF)** |  |  |  |  |  |  |  |  |  |  |  |
| No response | 49 | 30 (61.2%) | 16<br>(32.65%) | 3 (6.1%) | **29**<br>**(59.2%)** | 1 (2%) | 1 (2%) | 16<br>(32.6%) | 2<br>(4.1%) | 3 (6.1%) | 46 (93.9%) |
| Less than 100,000 | 120 | 71 (59.2%) | 43 (35.8%) | 6 (5%) | 38<br>(31.7%) | 8<br>(6.7%) | 0 (0%) | 63<br>(52.5%) | 11<br>(9.2%) | 1 (0.8%) | 119 (99.2%) |
| 100,000 - 199,000 | 24 | 8 (33.3%) | **16 (66.7%)** | 0 (0%) | 2 (8.3%) | 2<br>(8.3%) | 2<br>(8.3%) | 14<br>(58.3%) | 4<br>(16.7%) | 1 (4.2%) | 23 (95.8%) |
| 200,000 - 299,000 | 5 | 3 (60%) | 2 (40%) | 0 (0%) | 1 (20%) | 0 (0%) | 0 (0%) | 2 (40%) | 2 (40%) | 0 (0%) | 5 (100%) |
| 300,000 - 399,000 | 1 | 0 (0%) | **1 (100%)** | 0 (0%) | 0 (0%) | 0 (0%) | 0 (0%) | 1 (100%) | 0 (0%) | 0 (0%) | 1 (100%) |
| More than 400,000 | 2 | 0 (0%) | **2 (100%)** | 0 (0%) | 0 (0%) | 0 (0%) | 0 (0%) | 2 (100%) | 0 (0%) | 0 (0%) | 2 (100%) |

Fig 2 shows detailed frequencies of answers to questions regarding knowledge of FGS. While schistosomiasis was known by 91% of the respondents, only 45% had heard about FGS. More than half of the respondents recognized lower abdominal pain as the sign and symptom of FGS (70.6%), reduced fertility (75.1%) and miscarriage (56.2%) as the common complications of FGS, contact with contaminated water as increasing the risk of FGS infection (57.7%) and PZQ (Biltricide®) as the drug that treats symptoms related to FGS (50.2%). Similarly, 86.1% of the respondents mentioned that FGS can be spread through defecating and urinating in water by an infected person and treating all the infected persons (72.5%) as ways of preventing FGS by the respondents. For other questions, correct answers were found by less than 50% of community women.

**Fig 2.**
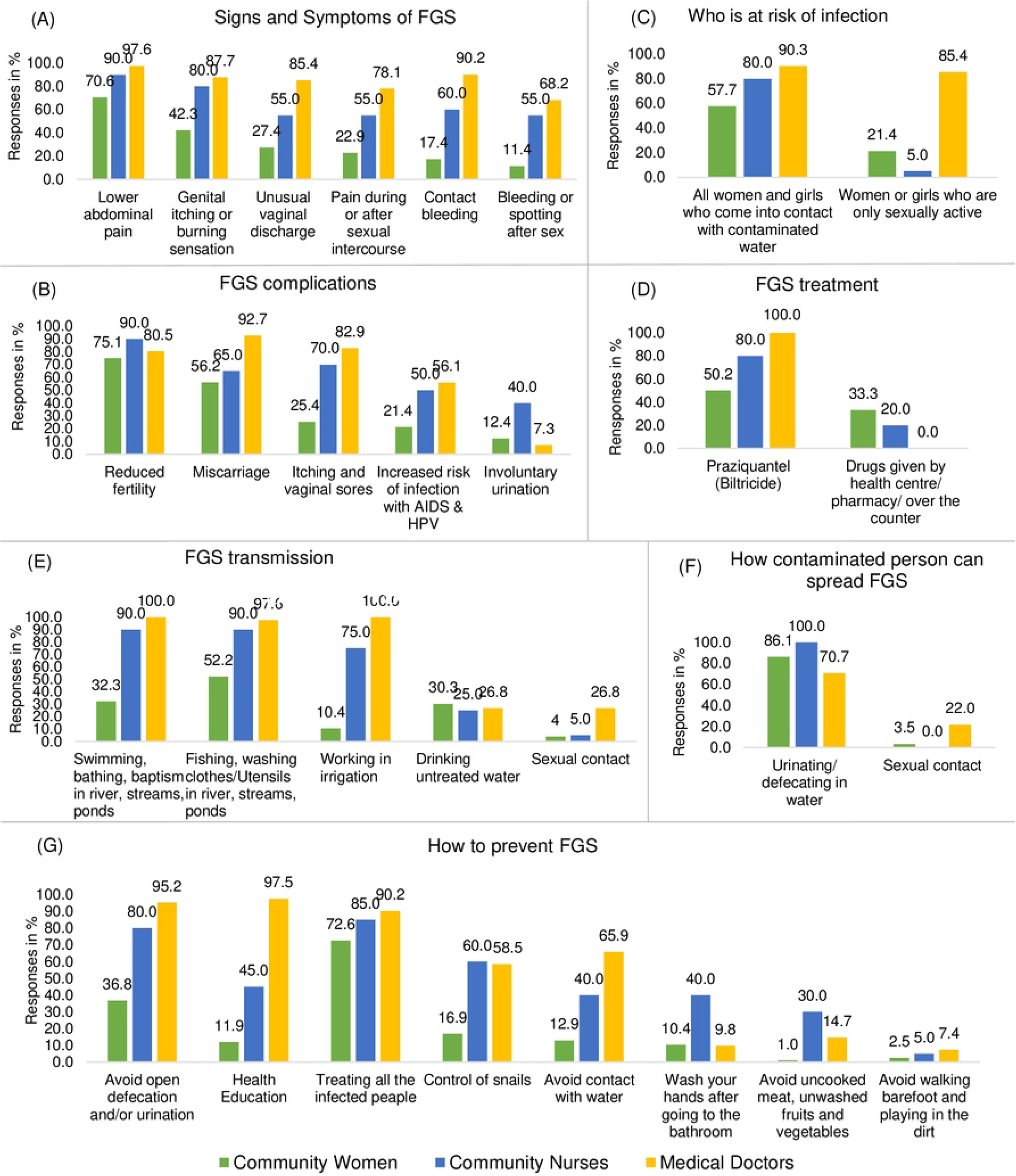
Community women’s, community nurses’ and medical doctors’ knowledge of female genital schistosomiasis in Kifua II and Kimpese, DRC.

Table 2 shows the association between community women’s knowledge on FGS and their sociodemographic characteristics. Age was significantly associated with knowledge of FGS transmission (φ=0.556, p=0.000), the group at risk of FGS (φ=0.521, p=0.04), and FGS disease prevention (0.518, p=0.000). Marital status was significantly associated with knowledge of signs and symptoms (φ=0.796, p=0.001), transmission mode (φ=0.595, p=0.001), and who is at risk of getting FGS (φ=0.298, p=0.016). Monthly income was significantly associated with knowledge on signs and symptoms (φ=0.866, p=0.000), FGS complications (φ=0.84, p=0.000), who is at risk of getting infected (0.322, p=0.002), and FGS prevention (φ=0.056, p=0.000). Type of occupation was significantly associated with knowledge on transmission mode (φ=0.669, p=0.000), who is at risk of getting infected (φ=0.344, p=0.000), FGS treatment (φ=0.313, p=0.000), and FGS prevention (φ=0.546, p=0.019). Level of education was significantly associated with knowledge on treatment (φ=0.224, p=0.02).

**Table 2.** Association between community women’s knowledge on female genital schistosomiasis and their sociodemographic background.

| Statement | Age |  | Marital status |  | Education level |  | Occupation type |  | Month income |  |
| --- | --- | --- | --- | --- | --- | --- | --- | --- | --- | --- |
| | $\phi$ | p-value | $\phi$ | p-value | $\phi$ | p-value | $\phi$ | p-value | $\phi$ | p-value |
| Signs and symptoms | 0.737 | 0.283 | **0.796** | **0.001\*** | 0.730 | 0.546 | 0.757 | 0.160 | **0.866** | **0.000\*** |
| Complications | 0.658 | 0.487 | 0.661 | 0.439 | 0.644 | 0.742 | 0.677 | 0.214 | **0.840** | **0.000\*** |
| Transmission | **0.556** | **0.000\*** | **0.595** | **0.001\*** | 0.517 | 0.687 | **0.699** | **0.000\*** | 0.512 | 0.752 |
| Who is at risk of infection | **0.521** | **0.004\*** | **0.298** | **0.016\*** | 0.228 | 0.895 | **0.344** | **0.000\*** | **0.322** | **0.002\*** |
| How infected persons spread FGS | 0.502 | 0.221 | **0.232** | **0.009\*** | 0.165 | 0.592 | 0.125 | 0.708 | **0.206** | **0.026\*** |
| Treatment | 0.461 | 0.811 | 0.199 | 0.130 | **0.224** | **0.020\*** | **0.313** | **0.000\*** | 0.194 | 0.064 |
| Prevention | **0.518** | **0.000\*** | **0.559** | **0.001\*** | 0.491 | 0.549 | **0.546** | **0.019\*** | **0.656** | **0.000\*** |

\*Cramer's V coefficient ( $\phi$ ) test of association was significant at $p<0.05$

#### 3.2.2 Healthcare professionals’ knowledge

Lower abdominal pain, contact bleeding and genital itching or burning sensation were the most (97.6%, 90.2% and 87.7% respectively) mentioned signs and symptoms of FGS by the doctors. Similarly, the nurses mentioned lower abdominal pain (90.0%) and genital itching or burning sensation (80.0%) as the common symptoms of FGS. Almost two-thirds (65.0%) of the nurses stated irregular menstruation as a sign of FGS, while only 17.1% of the doctors mentioned it (Fig 2A). Most (92.7%) of the doctors mentioned miscarriage as the complication of FGS, while reduced fertility was mentioned as the common complication by most (80.0%) of the nurses. Urinary incontinence (involuntary urination) was mentioned by 40.0% of the nurses and only 7.3% of the doctors (Fig 2B). The majority (85.4%) of the doctors believe that women or girls who are sexually active are at risk of acquiring FGS (Fig 2C). All the doctors and 80.0% of the nurses mentioned that FGS is treated by PZQ (biltricide) (Fig 2D). The doctors and the nurses had high knowledge of FGS transmission modes; however, few misconceptions were mentioned. These include drinking untreated water (doctors: 26.8% and nurses: 5.0%), sexual contact (doctors: 26.8% and nurses: 5.0%) and eating unwashed fruits and vegetables (nurses: 20.0% and doctors: 4.9%) (Fig 2E). Almost a third (29.3%) of the doctors did not mention defecating and urinating in water as how an infected person contributes to the spread of FGS (Fig 2F). For FGS prevention, health education was mentioned by almost all the doctors (97.5%) and by only 45% of the nurses. Avoiding open defecation/urination was said by 95.2% of the doctors and 80.0% of the nurses, while treating all the infected persons by 90.2% of the doctors and 85.0% of the nurses. Lastly, some misconceptions exist on ways to prevent FGS; including avoiding eating uncooked meat and unwashed fruits and vegetables (mentioned by 30.0% of the nurses and 14.7% of the doctors), washing hands after going to the toilet (mentioned by 40.0% of the nurses and 9.8% doctors), and avoiding walking barefoot and playing in the soil (mentioned by 7.4% of the doctors and 5.0% of the nurses) (Fig 2G).

### 3.2 Attitudes toward FGS

#### 3.2.1 Community women’s attitude

The Cronbach’s α coefficient was 0.934, indicating that the Likert scales were reliable and internally consistent. The means of the scores of attitude by sociodemographic characteristics of community women are shown in Table 1. Overall, more than half of them had a positive attitude, with 48.8% reaching a mean of 4 and 9.4% a mean of 5 (the most positive attitude). Almost one third of women (34.8%) had negative attitude towards FGS (mean = 1). Positive attitude (mean = 4) was the most frequent among all categories of sociodemographic characteristics, except among women aged 26-36 years, those with secondary and higher education, and those unemployed, with respectively 51.9%, 45.5%, 43% scoring a mean of 1.

Detailed responses to statements related to community women’s attitudes toward FGS are summarized in the Fig 3. About a third of women (34.8%) considered FGS not a very serious disease. Regarding disease diagnosis, 34.8% would not go to the hospital if infected and 35.3% would feel uncomfortable during gynaecological examination. As for treatment and prevention, 34.8% of women did not think taking FGS medication was important for their health, 72.1% found it difficult to avoid risky water contact and 41.3% didn’t think that defecating and urinating in the toilet was important for their health. Regarding the need of social support, 38.3% didn’t intend to share their FGS status with their husband nor their loved ones (63.6%), and 31.8% believed that their husbands would leave them if they were infertile.

**Fig 3.**
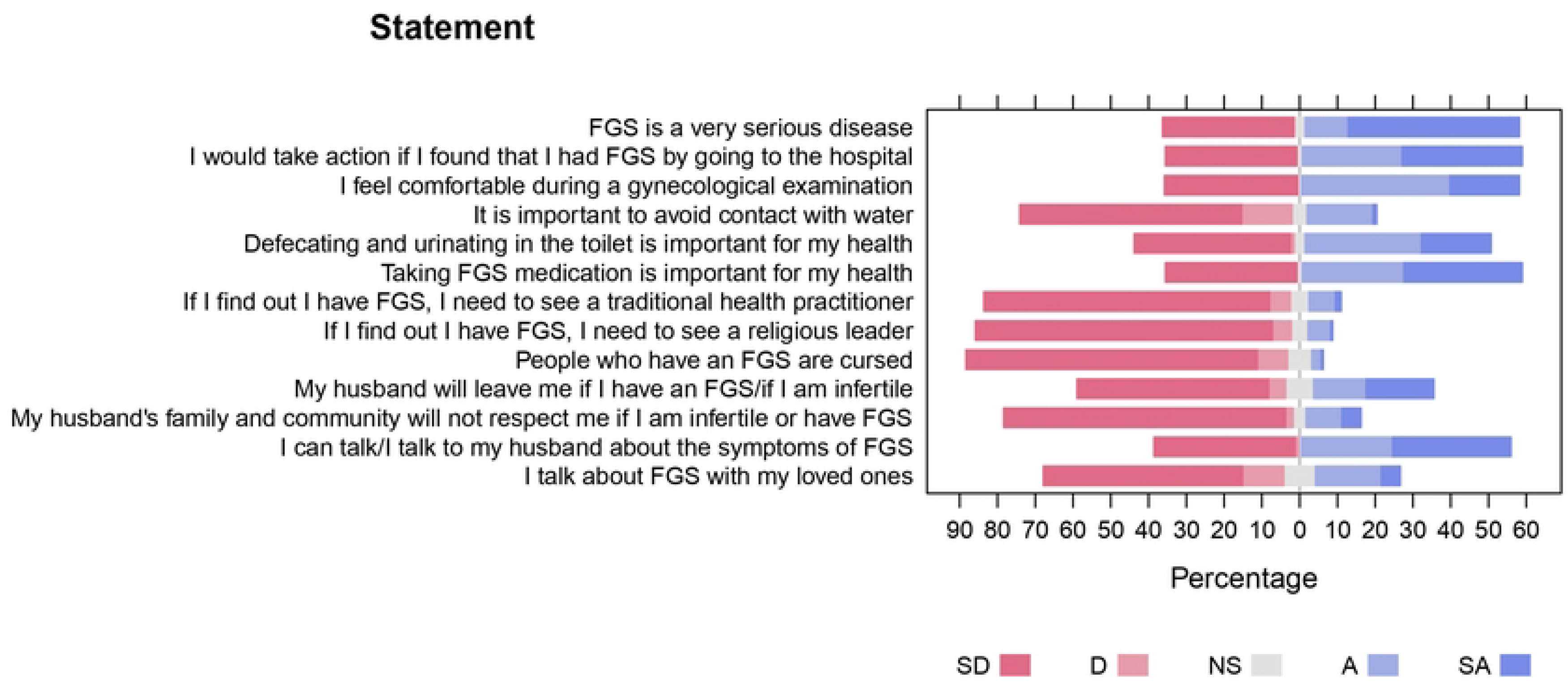
Community women’s attitudes toward female genital schistosomiasis in Kifua II, DRC (SD - Strongly Disagree; D - Disagree; NS - Not Sure; A - Agree; SA - Strongly Agree)

Associations between community women’s attitudes toward FGS and their socio-demographic characteristics are summarized in Table 3. Age and monthly income were found to have a positive association with how serious FGS is considered (γ=0.218, p=0.016; γ=0.244, p=0.025), openness to undergo gynaecological examination (γ=0.182, p=0.042; γ=0.342, p=0.001) and taking medication (γ=0.192, p=0.007; γ=0.24, p=0.017). This implies that younger women and those with lower monthly income do not believe that FGS is a serious disease, are uncomfortable to find out if they have FGS and do not agree that taking medication is important. Avoiding risky water contact had a significant and positive association with age (γ=0.312, p=0.001). Health importance of defecating and urinating in the toilet had a positive association with the level of income (γ=0.304, p=0.003) and occupation (γ=0.15.327, p=0.002). This implies that women with lower income do not consider avoiding open defecation or urination unhygienic or important for their health. Visiting a traditional healer was found to have a positive and significant association with age (γ=0.386, p=0.003), monthly income (γ=0.375, p=0.006) and type of occupation (γ=23.33 p=0.000). This suggests that older women with more income would not visit a traditional healer if they found out they have FGS. Talking to their husbands or loved ones regarding FGS symptoms was positively associated with monthly income (γ=0.214, p=0.034; γ=0.26, p=0.012) and occupation (γ=9.354, p=0.025; γ=17.818, p=0.000). These infer that community women with lower income are less willing to open up and talk about their symptoms to other people.

**Table 3.**
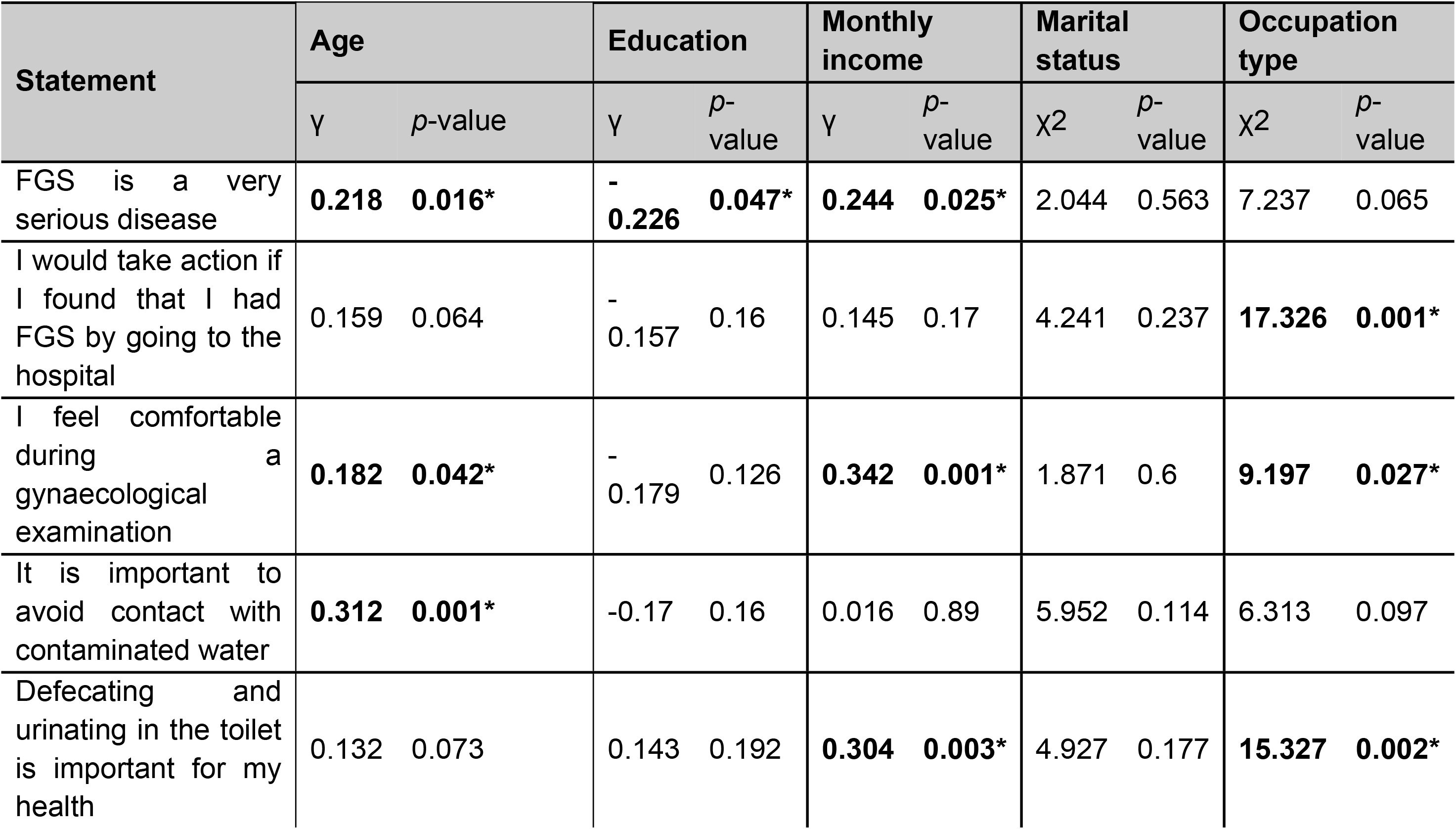
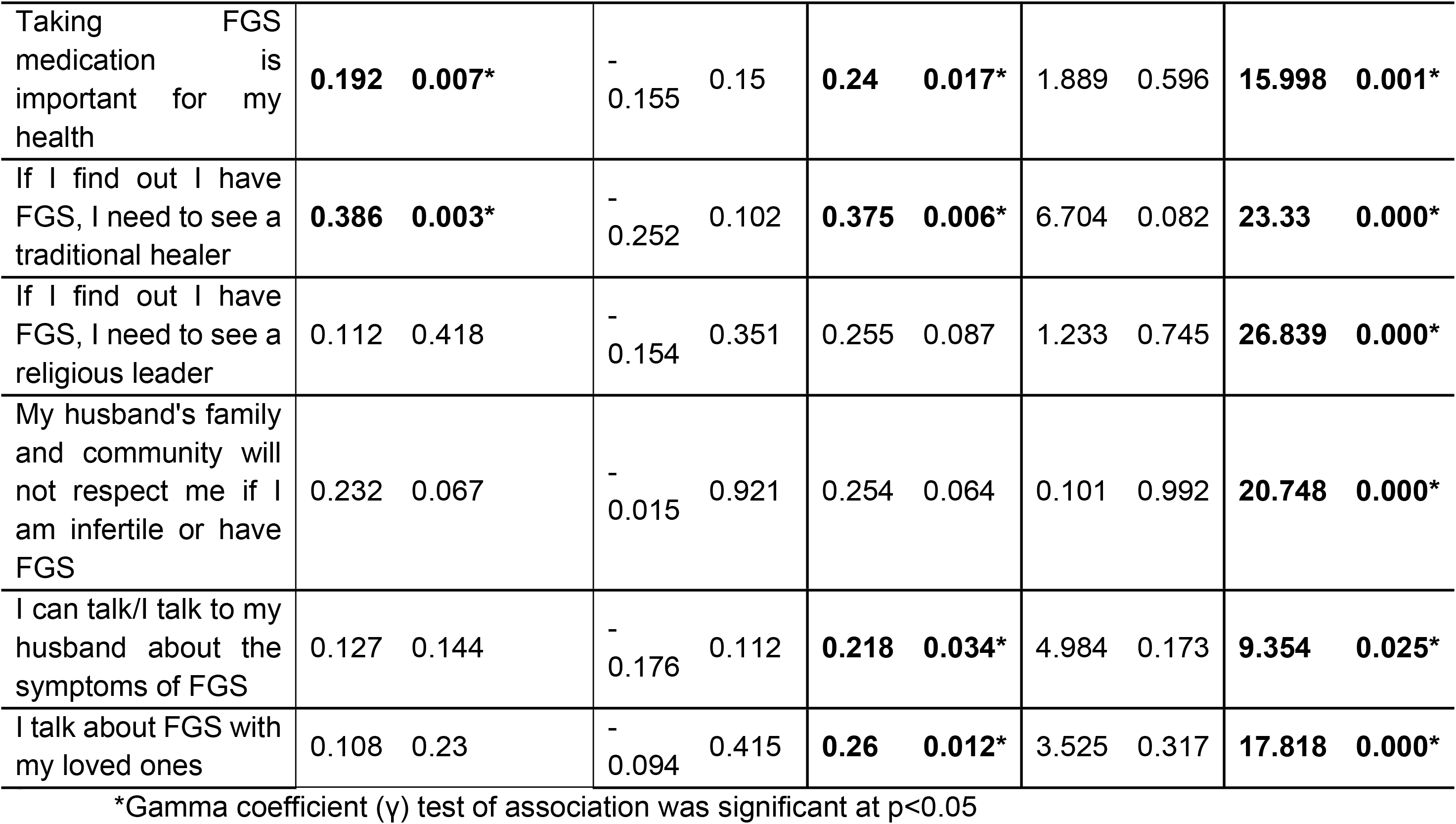
Association between community women’s attitudes toward FGS and their socio-demographic background.

#### 3.2.2 Healthcare professionals’ attitude

Detailed responses to statements related to healthcare professionals’ attitudes toward FGS are shown in the Fig 4. Most (81.9%) of the healthcare professionals regard FGS a very serious disease and 95.7% believe that it is necessary to advocate for its prevention measures. Similarly, 96.7% consider diagnosing FGS their responsibility and very important for them. When asked if the medication for FGS is expensive, 62.3% were not sure, while 18.0% disagreed and 11.5% agreed that FGS medication is expensive. Most (91.8%) of the respondents agreed to talk about FGS to their colleagues, while only 6.6% were not sure if they could.

**Fig 4.**
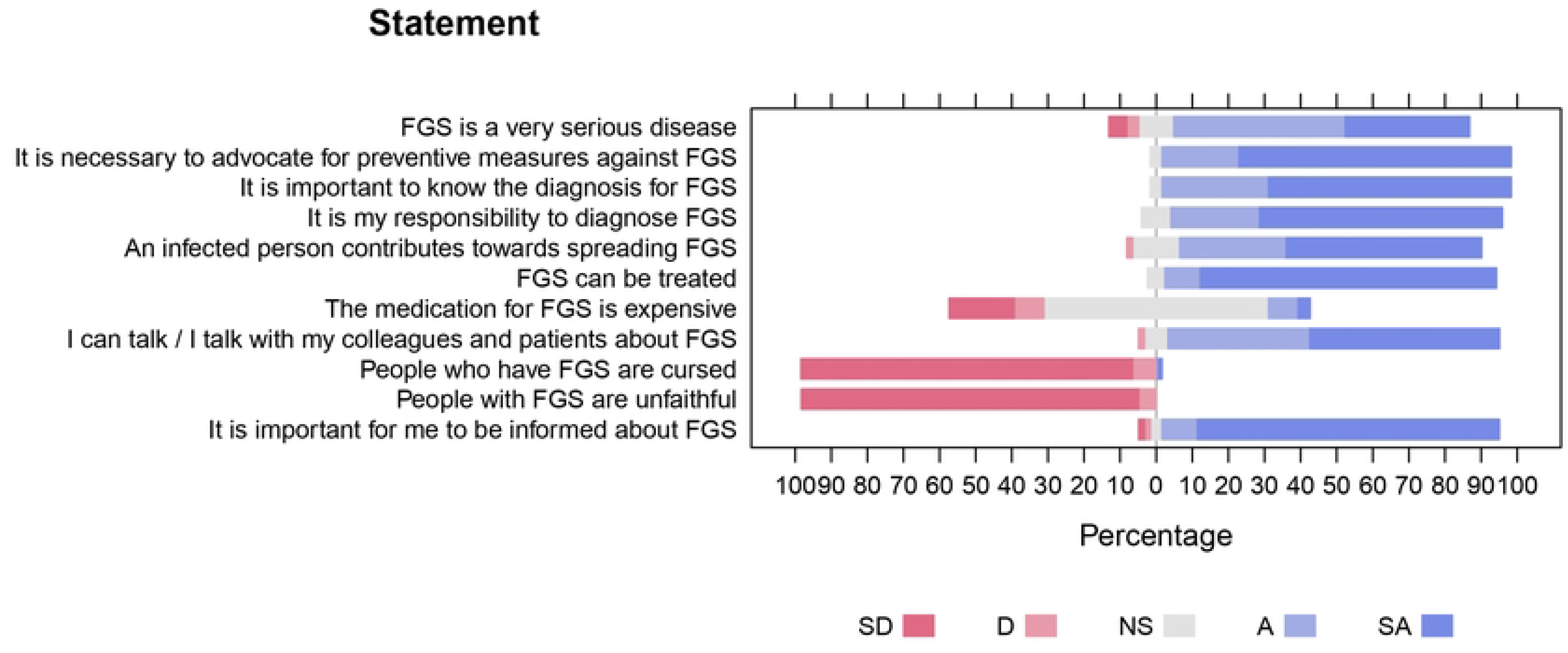
Healthcare professionals’ attitudes and perception toward female genital schistosomiasis.

### 3.3 Practices

#### 3.3.1 Community women practices

##### a) Water, sanitation, and hygiene practices

As shown in Table 4, the main source of water for household use was protected wells (42.3%) or river/stream (40.3%). More than three-quarters (77.6%) of the respondents come into contact with the river/stream daily through fetching water, followed by agriculture (48.3%) cassava retting (43.3%), washing dishes and laundry (38.3%) and bathing (31.1%). Almost two-thirds (63.2%) visit the river/stream twice a day, and about 50% spend more than 15 minutes in the water. At least 31.3% of the respondents do not own a latrine, while a large majority (70.6%) defecate or urinate outside the toilet, mostly in the bush (59.7%), because they do not have access to a toilet (46.8%). This happens on a daily basis for 46.8% of the respondents.

**Table 4.** Summary of community women’s practices, water, sanitation, and hygiene regarding FGS.

| Variable | Response | Frequency | Percentage |
| --- | --- | --- | --- |
| Main source of water for the households | Protected well | 85 | 42.3 |
|  | River /stream | 81 | 40.3 |
|  | Piped water | 33 | 16.4 |
|  | Unprotected well | 2 | 1.0 |
| Activities leading to water contact | Fetch water | 156 | 77.6 |
|  | Agriculture | 97 | 48.3 |
|  | Cassava retting | 87 | 43.3 |
|  | Laundry & washing dishes | 77 | 38.3 |
|  | Bathing | 63 | 31.3 |
|  | Fishing | 28 | 13.9 |
|  | Baptism | 10 | 5.0 |
|  | Swimming | 8 | 4.0 |
| Time taken for activities in contact with water | More than 15 minutes | 101 | 50.2 |
|  | Less than 5 minutes | 56 | 27.9 |
|  | 5 to 10 minutes | 25 | 12.4 |
|  | 10 to 15 minutes | 19 | 9.5 |
| Number of times for daily contact with water | Twice | 127 | 63.2 |
|  | Once | 46 | 22.9 |
|  | More than 3 times | 17 | 8.5 |
|  | Thrice | 11 | 5.5 |
| Type of toilet for the household | Pit latrine | 89 | 44.3 |
|  | No toilet | 63 | 31.3 |
|  | Communal latrine | 39 | 19.4 |
|  | Flush toilet | 10 | 5.0 |
| If defecate/urinate outside the toilet | Yes | 142 | 70.6 |
|  | No | 59 | 29.4 |
| Where defecated/urinated outside the toilet | In the bush | 120 | 59.7 |
|  | In an open space | 65 | 32.3 |
|  | In the water | 22 | 10.9 |
|  | In a tin bucket | 5 | 2.5 |
|  | I don't remember. | 2 | 1.0 |
| Why did not use the toilet | No access to the toilet | 94 | 46.8 |
|  | The Call of Nature | 37 | 18.4 |
|  | The toilet was busy/cluttered | 18 | 9.0 |
|  | The toilet is there but poorly maintained | 14 | 7.0 |
|  | When in the field | 9 | 4.5 |
|  | I don't like using the toilet | 5 | 2.5 |
|  | Lack of water to supply the toilet | 1 | 0.5 |
|  | The toilet is far from the house | 1 | 0.5 |
| How many times of defecation/urination outside the toilet | Every day and when nature calls | 87 | 43.3 |
|  | I do not remember | 42 | 20.9 |
|  | It was only once | 10 | 5.0 |
|  | Occasionally and when in the field | 3 | 1.5 |

Associations between socio-cultural factors and WASH practices of the community women and their socio-demographic characteristics are shown in Table 5. Age (φ=0.664, p=0.000), marital status (φ=0.7, p=0.000), and monthly income (φ=0.682, p=0.007) were found to have a statistically significant association with activities done in water. Level of education (φ=0.239, p=0.012) was associated with the type of toilet used in the household. Defecating or urinating outside the toilet was found to be associated with age, marital status, level of education and occupation. Lastly, type of occupation (φ=0.348, p=0.051), had a statistically significant relationship with the action taken when symptoms are experienced (visiting a health facility or not).

**Table 5.**
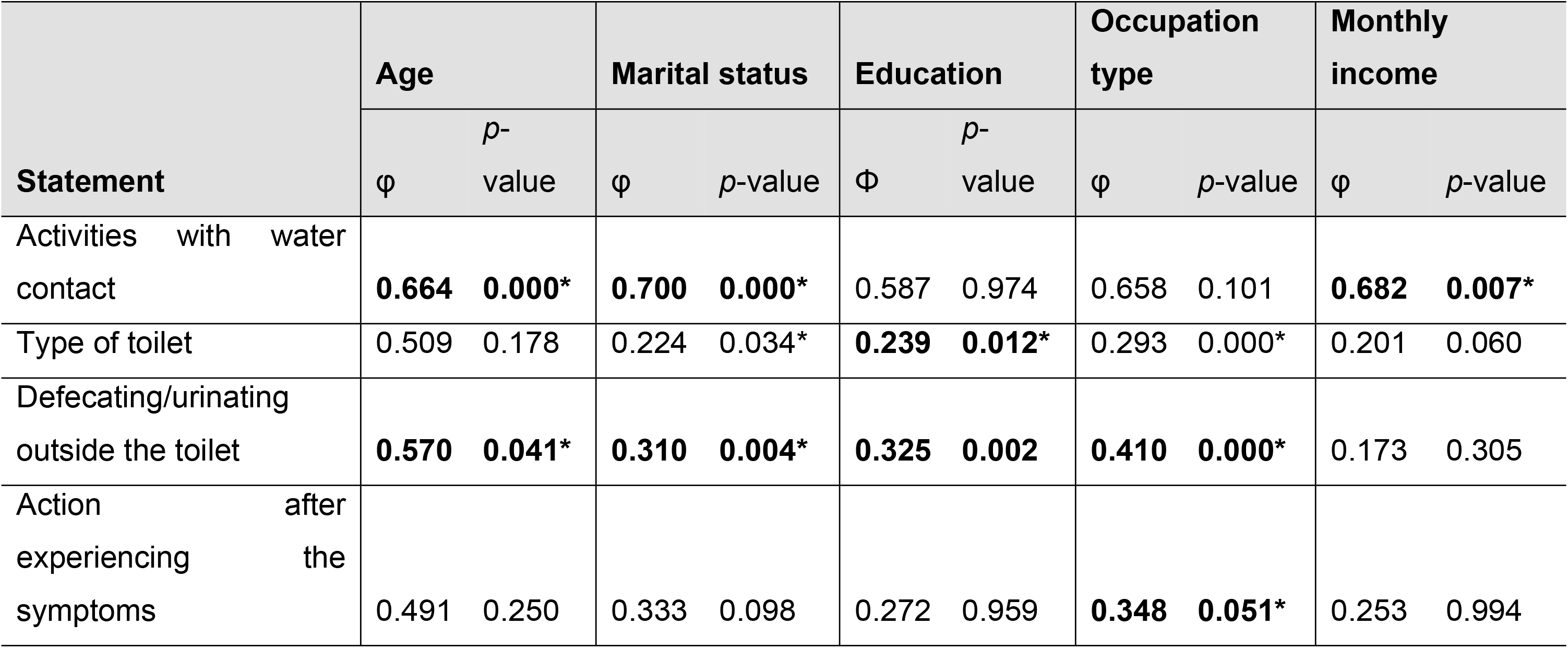
Association between socio-cultural factors and WASH practices of the community women and their socio-demographic characteristics.

##### b) Community women health-seeking behaviour

Nearly three-quarters (73.1%) of the respondents stated that they would / visit the health facility when they experienced / will experience any of the symptoms related to FGS. Some (15.0%) stated that they would do nothing. Urine and stool tests were mentioned by 52.2% and 34.3% of the respondents respectively, as the tests done when FGS related symptoms are experienced. More than a half (57.7%) of the respondents reported to have received PZQ/biltricide via a treatment campaign, while a few (8.0%) did not because they did not know while family and friends advised against the treatment for 5.0%. Nearly all (95.0%) the respondents stated that they would accept a vaccine if available (Table 6).

**Table 6.**
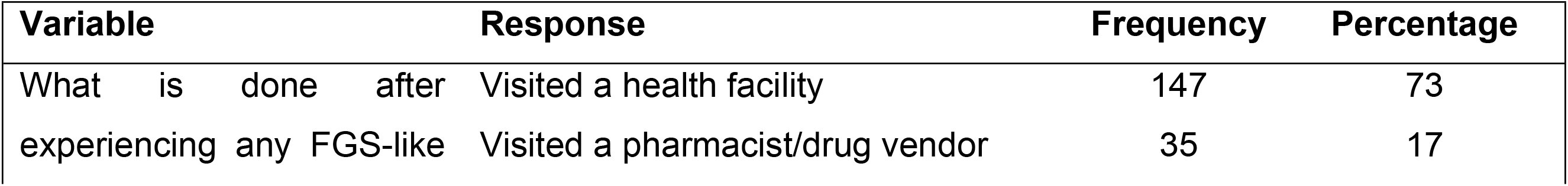
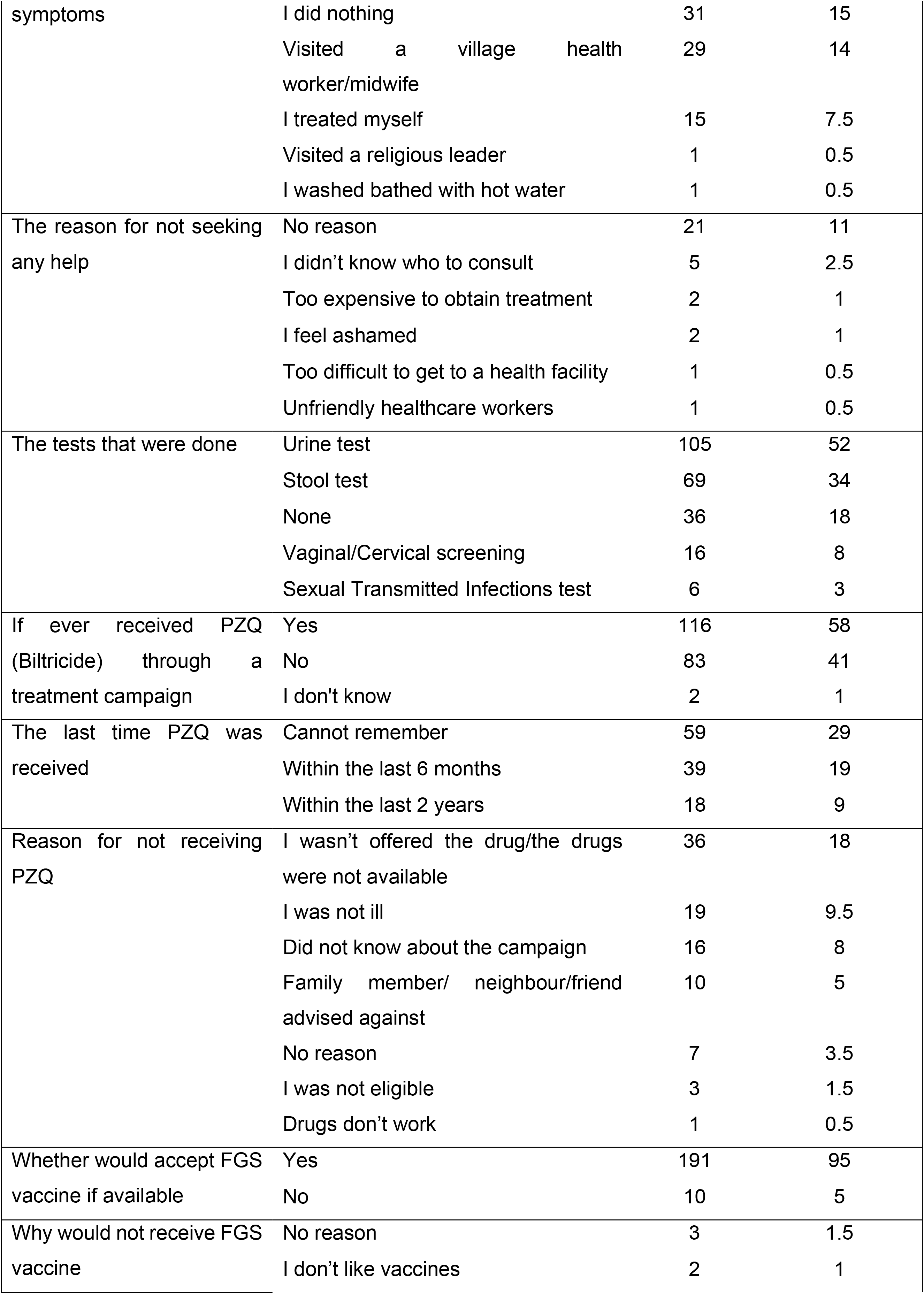
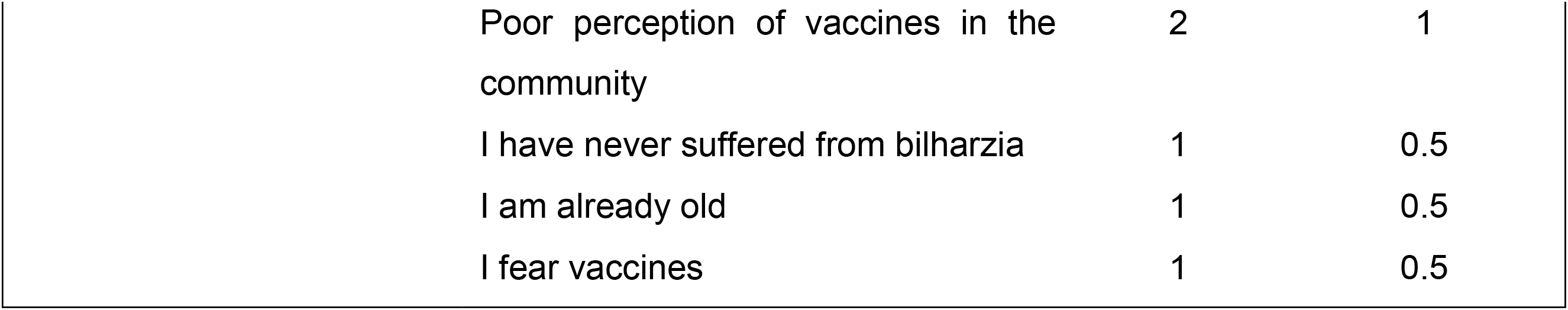
Summary of community women’s health-seeking behaviour in Kifua II.

#### 3.3.2 Healthcare professionals’ practices and management of FGS

More than three-quarters (77%) of the respondents consider FGS prevalent in Kimpese and 65.6% of them mentioned having had patients with FGS. Some (9.8%) respondents stated to have between 5-25 cases and 8.2% between 25 -35 cases. Of those who had patients with FGS, when asked how they diagnosed the patients, 21% mentioned by clinical examination of urine sediments and stool, while 4% via symptomology. Another diagnosis mentioned was vaginal smear to reveal the *Schistosoma* species eggs. For FGS diagnostic test (referring to a microscope), 57.4% of the respondents mentioned they are available in their working stations and only 49.2% had sometimes access to PZQ (SI6).

## 4. DISCUSSION

This study assessed, for the first time, the KAP toward FGS among community women and health professionals in a well-known endemic area of the Kongo Central province of DRC [24,26]. Such knowledge could help in designing tailored control strategies. While 93% of women claimed to know about schistosomiasis, more than half (55%) had a low level of knowledge about FGS. This is in line with other studies conducted in endemic sub-Saharan African settings, which reported a higher level of knowledge about urinary schistosomiasis compared to the knowledge of FGS [32–34]. Higher knowledge of urinary schistosomiasis can be explained by the fact that urinary schistosomiasis is generally well recognized by the presence of blood in urine, which is a sensitive and specific symptom. In addition, this might be result of long-standing endemicity, contact with research teams and/or the organization of schistosomiasis control campaigns, through mass treatment or health education. In our study area schistosomiasis is well known since Lengeler at al successfully assessed the use of questionnaires on urinary schistosomiasis to map both intestinal and urinary schistosomiasis [35]. Awareness about schistosomiasis could also increase through sensitization activities conducted in the framework of research studies or through MDA campaigns which took place recently in the region [24,25,36].

The association between high income and high knowledge of FGS symptoms in this study could partly be explained by the fact that people with high income are more likely to access information from sources such as written material or radio and television. A similar observation was made in endemic communities surrounding Lake Albert in Uganda [28].

As expected, medical doctors and nurses had a higher knowledge of FGS symptoms than the community women As reported previously by Linsuke et al. [36], healthcare staff in this region are aware of schistosomiasis symptoms, including haematuria as an indication of *S. haematobium*. This is probably linked to regular exposure to cases and the region’s long history of high schistosomiasis prevalence [26,36].

Our study recorded some misconceptions, mainly linked to FGS transmission, with one in four (27%) medical doctors linking FGS transmission to sexual contact, and 85% clamming that only women and girls who are sexually active are at risk of FGS infection. The same responses were given by 4% and 21% of the community women, respectively. Similar misconception of sexual transmission of FGS has been reported in studies conducted among local health workers in Zanzibar and Ghana [37,38]. These misconceptions could be linked to confusion, as reported in Ghana, Tanzania and Zanzibar, where the respondents could not clearly distinguish the transmission modes from those of other water-borne diseases and sexually transmitted infections (STI) [32,33,39]. Moreover, only half of the health professionals linked FGS with an increased risk of infection with HIV and HPV. These results clearly point the need for updating knowledge of health professionals regarding FGS. In this context, WHO FGS training guides and atlas for primary healthcare workers to manage FGS in limited-resource settings in endemic regions could be helpful. Other misconceptions included one in four health professionals and one in three of the community women who mentioned drinking untreated water and eating unwashed fruits and vegetables as transmission modes for FGS. Also, only a third of the community women linked transmission of FGS with swimming in the rivers, ponds, and streams. This was similar to what was found in the local community of Mwea in Kenya (33%) [23], but much lower than the proportion reported in the communities living around Lake Albert, Uganda (81%) [28].

The negative attitude that open defecation/urination is not unhygienic (41%) might be attributed to not owning a latrine as only 49% have a latrine in their household (see below). An alternative hypothesis is that open defecation and urination have been normalised as it has been going on for a long time [40]. Furthermore, some of the women do not consider FGS a serious disease; thus, knowing whether they have FGS or taking medication was not so important to them. This is an important local risk factor contributing to disease persistence in the region. Many women (72%), especially the younger ones, do not consider avoiding contact with contaminated water sources important. This sentiment is possibly linked to the fact that the Kifua II community depends directly on the water bodies for their livelihood and water needs. Also, other studies have shown that younger women and girls are the ones who tend to conduct most household chores, such as collecting water, and washing utensils or clothes, which inevitably makes it difficult to avoid water contact [28,41]. On the other hand, talking about FGS-like symptoms is considered a taboo, as women do not speak freely and 38% would not share their experiences and symptoms with their husbands, and 64% not even with their loved ones. This is also related to the belief that they would be left by their husbands or that their husbands would find a second wife if they fail to bear children (31.8%). Moreover, the cultural burden that tags the woman’s value on how many children she bears worsens this [42].

Although the healthcare professionals recognise that FGS is a serious disease and that diagnosis of FGS is their responsibility, user friendly diagnostic technologies are limited (only 57.4% of the healthcare workers had a microscope in their facility) and this is worsened by the few skilled personnel. This is in line with the survey by Linsuke et al. [36] who reported that only 42% of the hospitals in Kongo Central perform urine sedimentation to diagnose *S. haematobium*, while none of the hospitals performs urine filtration. Therefore, most low-resource health facilities in DRC depend on syndromic approaches for diagnosis [36,43]. A cross-sectional study conducted in a nearby Health Zone (Kisantu) reported the presence of urinary schistosomiasis among pregnant women (17.4%), ranging from asymptomatic to low symptomology [43]. Furthermore, their findings confirmed coinfections of urinary schistosomiasis and STIs (*Chlamydia trachomatis*, *Neisseria gonorrhoeae* and *Trichomonas vaginalis*) [43]. Therefore, relying on syndromic management alone cannot suffice, as medication for the different conditions differ despite the shared symptoms. For instance, FGS is treated by anthelminthic drugs, while STIs are treated by antibacterial or antiviral drugs. Moreover, the lack of equipment leads to underdiagnosis [9,38]. Apart from microscopes, clinicians need specialised equipment like colposcopy, and specialised training to gain competence in FGS diagnosis.

While the village has few boreholes, obtaining water from them is not free of charge and this is costly for households with low income (more than half earn < USD 50 monthly). Consequently, a significant number of women (77.6%) undertake risky water practices daily, despite being aware of the risk of infection. Some households only purchase water for cooking and drinking, while the rivers and streams provide water for other domestic activities such as washing clothes and dishes (38.3%), bathing (31.3%) and cassava retting (43.3%). In 2010, Onyeneho [44] made similar observations in Nigeria, where community members opted for water from contaminated sources because they were nearby and the boreholes were quite distant. The lack of latrines in a third of the respondent households could explain the rampant open defecation /urination. In the last health survey report released for 2013-2014, more than 20% of rural households in DRC did not have access to any sanitation facility [45]. In our study area (Kifua II), 31.3% of the households did not have a toilet/latrine, and 70.6% of the respondents urinated/defecated outside the toilet. This huge sanitation problem contributes greatly to disease transmission. In addition, working in agricultural fields for many hours without latrines contributes to this. These fields are usually also located close to the water bodies (observed during the fieldwork), and thus the excreta is washed down into the water bodies when it rains. Their negative attitude that it is not unhygienic to defecate and / or urinate outside the toilet is also likely to contribute to this behaviour. Anyolitho and colleagues [28] reported that people in Ugandan communities surrounding Lake Albert believe that not defecating in the lake compromises their fish yield. In a study conducted among female heads of households in Mushandike Resettlement Irrigation Scheme, Zimbabwe, they reported 100% use of the constructed field toilets whilst working in the fields [46]. The toilets were built in a layout scheme that ensured people are nearer to them at all times than to a bush, hence promoting their usage. There is a need to consider building field latrines in Kifua II so that people working in the fields are always closer to the latrines [47]. Moreover, providing piped water supply at a central point at a reduced cost or adding the number of boreholes and washing slabs could also be considered to reduce the use of natural water bodies. Increasing the use of toilets and boreholes could considerably reduce schistosomiasis (hence, FGS) in the communities.

Studies have reported cases where women and girls experiencing FGS-like signs and symptoms face stigmatization, thereby negatively impacting their health-seeking behaviour [33,48,49]. However, stigma is generally low in Kifua II, probably because the community has a considerably high knowledge of schistosomiasis. It should be noted that this village has participated in many schistosomiasis-related studies before the start of this survey as mentioned above. This is probably the reason why our findings differ from other FGS KAP surveys conducted in several parts of Africa, reporting that infected and affected women or girls suffer both socially and mentally [38,50]. In some communities, young girls and women were labelled as “prostitutes” for presenting with FGS or STIs symptoms [33,38]. These stereotypes may hinder them to seek medical attention for the fear of being regarded as promiscuous. Additionally, Anyolitho et al. [28] reported that people who suffer from chronic effects of schistosomiasis such as swollen bellies and emaciation suffer from high stigma.

However, to thoroughly evaluate self and external stigma among community women and girls in Kifua II, further surveys that include in-depth qualitative interviews and multiple focus group discussions need to be carried out. This is because a KAP survey by interviews only, records individual opinions based on the statement given. There might be considerable gaps in what is said and what is done [51], especially around sensitive cultural issues normally not spoken in public.

It is encouraging that the acceptability of the treatment is high in Kifua II, and most of the women (95%) mentioned they would accept a schistosomiasis vaccine if made available. Similar reports of high treatment acceptability were obtained in a survey in Mozambique [52], while this is not the case in the north-east and south-west of Uganda, where there is persistent fear to take treatment because of conspiracy theories and rumours behind the ‘real’ objective of the MDA campaigns and subjective side effects of the drugs [28,52,53]. In Morogoro, Tanzania, MDA for schistosomiasis were rejected through community riots, due to the failure of communication between the campaign programs’ leaders and the local people [53]. Rumours spread that these campaigns are part of the government’s covert birth control campaigns, while others claimed that the pills’ side effects are fatal [53,54].

## CONCLUSION

Community women and the health professionals of the Kimpese region are well aware of schistosomiasis, but FGS-specific knowledge is low and misconceptions about disease transmission are evident. Besides, most women still undertake risky water contact practices and do not use latrines, thus predisposing themselves and the community to FGS. Extensive qualitative research is needed to better understand factors underlying the variability of KAP among community women, and why the community engages in risky behaviour and fail to adhere to preventive measures despite being aware. This will help to optimise or adapt the WASH interventions and health communication. It is also crucial for healthcare professionals to acquire the right competencies regarding FGS as they are at the front line to offer primary health care and counselling on FGS to the communities. We concur with other researchers in recommending the integration of FGS with other routine women’s reproductive health services as part of long-term control and preventive measures.

## Data Availability

All the data is fully available and accessible in the main manuscript and supplementary information without any restrictions

## Author contributions

1. **Conceptualization:** Cecilia Wangari Wambui, Tine Huyse, Joule Madinga

2. **Data collection:** Cecilia Wangari Wambui

3. **Formal analysis:** Cecilia Wangari Wambui, Joule Madinga, Mercy Gloria Ashepet, Maxson Kenneth Anyolitho

4. **Methodology:** Joule Madinga, Cecilia Wangari Wambui, Maxson Anyolitho, Tine Huyse,

5. **Project administration and coordination:** Tine Huyse and Joule Madinga

6. **Supervision:** Tine Huyse, Mercy Gloria Ashepet and Joule Madinga

7. **Writing – original draft and final copy:** Cecilia Wangari Wambui and Joule Madinga,

8. **Writing – review & editing:** Tine Huyse, Mercy Gloria Ashepet and Maxson Kenneth Anyolitho

## Conflict of interest

We declare no conflicting interests whatsoever.

## Acknowledgment

This research is financed by the ATRAP project of the Development Cooperation program of the Royal Museum for Central Africa with support of the Directorate-General Development Cooperation and Humanitarian Aid. We would also like to thank the Kifua II community, the Kimpese healthcare professionals, the 8 field researchers (Edith Matota, Merveil Mafwana, Marceline Luntadila, Nana Nzuzi, Falone Nsimba, Gisele Nzuzi, Esther Miezi, and Benoir Nsimba) who aided in data collection, Germain Kapour, Théo Emboni and Chantal Mokoko for their support in the field. We also thank Dr. David De Coninck for his insights on data analysis. Gratitude goes to Stephen Ndung’u who assisted and guided the entry of the questionnaires into the KoboCollect toolbox and Pierre Sacré who assisted in translating the questionnaires from English to French and back to English.

## Notes

### Competing Interest Statement

The authors have declared no competing interest.

### Funding Statement

Yes

### Author Declarations

The study was approved by the Ethical Committee of the University of Kinshasa, Democratic Republic of Congo (no191/CNES/BN/PMMF/2020)

